# DASH diet, Reduced Rank Regression Dietary Patterns and relations with kidney function in the CHRIS general population study

**DOI:** 10.1101/2024.07.18.24310620

**Authors:** Giulia Barbieri, Vanessa Garcia-Larsen, Rebecca Lundin, Ryosuke Fujii, Pietro Manuel Ferraro, Giovanni Gambaro, Roberto Melotti, Martin Gögele, Lucia Cazzoletti, Peter P Pramstaller, Maria Elisabetta Zanolin, Cristian Pattaro, Essi Hantikainen

## Abstract

**Objective:** We assessed dietary patterns (DPs) using the pre-defined Dietary Approaches to Stop Hypertension (DASH) score and reduced rank regression (RRR) to evaluate their association with estimated glomerular filtration rate (eGFR) in a general population sample.

**Methods:** We analysed cross-sectional data from 6,133 healthy adult participants of the Cooperative Health Research In South Tyrol (CHRIS) study. Using self-reported food frequency questionnaire data, we derived the DASH-score and two RRR-based sex-specific DP-scores based on nine cardio-renal-metabolic parameters. We fitted sex-stratified linear and non-linear models to assess associations with creatinine-based eGFR.

**Results:** Males with higher intake of cereals, whole grains, sugar, fruits, and legumes, and lower intake of beer, and red and processed meat (MDP_1_), exhibited higher eGFR levels. Coherently, high adherence to the DASH-style diet was associated with higher eGFR. In females, the associations varied by menstrual status. Among those with ceased menstruation, a low intake of meat, spirits, refined grains, beer, and fish, and a high intake of whole grains and dairy products was associated with higher eGFR. In females still experiencing regular menstruation, a high intake of beef, nuts, beer, legumes, fish, and coffee, and low intake of refined grains, (FDP_2_) was associated with lower eGFR. No association was observed with the DASH score in females.

**Conclusion:** Our findings confirm the DASH-style diet is associated with better kidney function in males, but not in females. By identifying sex-specific kidney function-oriented DPs, the RRR approach provides new insights into the diet-eGFR relation, highlighting in addition potential effect modification by menstrual status.

## Introduction

Already affecting approximately 10% of the global adult population, chronic kidney disease (CKD) has become a major public health concern.[1] The increasing prevalence of chronic conditions like diabetes, cardiovascular disease (CVD), and hypertension, alongside population aging, are the most recognized factors contributing to the growing CKD burden.[2] Prevention is thus fundamental to reduce exposure to avoidable risk factors.[3]

Given the substantial overlap between CVD and CKD, lifestyle recommendations often coincide, particularly with regard to dietary habits. The Dietary Approach to Stop Hypertension (DASH) Diet has been specifically designed to prevent and manage high blood pressure by recommending a high intake of grains, fruits, vegetables, and low-fat dairy, while limiting the consumption of meat, fats, and sweets. General population studies suggest a protective association between adherence to DASH diet and risk of CKD onset and progression.[4, 5] The DASH diet has further been recommended by the National Institute of Diabetes and Digestive and Kidney Diseases as well as the National Kidney Foundation for non-dialytic CKD patients to improve kidney health.[6] Nevertheless, the pathophysiological role of diet and its components in kidney function is not yet fully ascertained. Further research is required to assess the relevance of these findings with regards to primary prevention of CKD.

The DASH diet excels in its established health benefits and broad applicability, allowing for consistent comparison across populations. However, in population studies, assessing adherence to predefined dietary indices may not always accurately capture specific dietary habits of individual populations as they consider only specific aspects of the diet, often leading to unclear interpretation of intermediate scores characterized by more complex nutritional compositions and dietary patterns (DPs).[7] In contrast, hybrid methods, such as Reduced Rank Regression (RRR), have emerged as useful exploratory approaches as they implement prior knowledge of the diet-disease pathway and allow identification of population-specific DPs, by considering the overall diet, associated with disease risk.[8, 9] Despite the increased complexity of interpretation and limited reproducibility in external studies, a hybrid approach offers flexibility in dietary pattern analysis compared to standard hypothesis-based dietary indices. This is particularly relevant in the context of CKD prevention, where ad hoc dietary scores have not yet been established and validated.

To date, only two studies have applied RRR in relation to kidney health using biomarkers as mediators. Cai et al. applied RRR to investigate associations between eGFR-based DPs and eGFR decline or incident CKD in a CKD-free large population-based cohort.[10] Kurniawan et al. derived RRR based DPs implementing a broader set of CKD-related risk biomarkers as mediators and investigated association with severity of impaired kidney function in population based study among participants with CKD.[11] Moreover, no formal comparison between DASH-like scores and RRR-like methods have been conducted. With the aim of gaining a complementary and comprehensive understanding of the role of diet and its association with eGFR, we compared the effects of the DASH diet score (DASH-score) with those of DPs specifically derived from RRR for risk factors of reduced estimated glomerular filtration rate (eGFR), the primary marker of kidney function. The analysis was conducted in a large and generally healthy population sample.

## Methods

### Study design

Analyses were conducted within the Cooperative Health Research in South Tyrol (CHRIS) study, a population-based study that recruited 13,393 adults from the Vinschgau/Val Venosta district (South Tyrol, Italy) between 2011 and 2018, as extensively described elsewhere.[11] Participants were invited to the study centre in the early morning following overnight fasting. They underwent blood drawing, urine collection, anthropometric analysis, blood pressure measurements, clinical examinations, and standardized self- and interviewer-administered questionnaires about medical history and lifestyle. The validated Global Allergy and Asthma Network of Excellence (GA^2^LEN) food frequency questionnaire (FFQ),[12] was introduced in 2014 and filled out by 8,842 participants.

We excluded participants reporting a diagnosis of any kidney disease,[13] hypertension, or diabetes, according to the questionnaire interview or the provision of corresponding medications (**Supplementary Table S1**). We further excluded 5 participants with >80% missing laboratory biomarker data, 34 participants with >20% missing FFQ items, and 86 out of the 0.5^th^-99.5^th^ percentile range of the total energy intake (TEI)/basal metabolic rate ratio (see flowchart in **Supplementary Figure S1**). Residual missing values in the FFQ were assumed to indicate non-consumption and imputed as ‘rarely or never’.[14] Missingness in all other variables was dealt with Multiple Imputations by Chained Equations (MICE) using the ‘mice’ R package v3.14.0.[15] Sample characteristics remained stable between the imputed and unimputed datasets. Eventually, 6,133 individuals were available for analysis.

### Outcome

The outcome of our analyses was eGFR, estimated from serum creatinine using the 2021 CKD-EPI equation, using the R package ‘nephro’ v1.3.[16]

### Assessment of Dietary Intake

Participants were asked to report in the GA^2^LEN FFQ how often they typically consumed one portion of each of 229 foods and beverages over the last year, selecting one among various options: rarely or never; once per month; once per week; 2-4 times per week; 5-6 times per week; once per day; and 2+ times per day. The consumption of each item was converted to grams per day, using the most recent edition of the Composition of Foods Table[17] to further determine macro- and micro-nutrients and total energy intake (TEI; kcal/day).

### Definition of the DASH-score

Adherence to a DASH-style diet was assessed based on eight different food components known for their beneficial or detrimental effects on the risk of hypertension, according to Fung et al.[18] The DASH-diet relevant FFQ-derived food groups (**Supplementary Table S2**), expressed in servings per week, were classified as ‘healthy’ (fruits, vegetables, whole grains, low-fat dairy products, and nuts and legumes) and ‘unhealthy’ (red and processed meats, sodium, and sugar-sweetened beverages (SSB)). The nutrient residual method was used to adjust the DASH components for sex-specific energy intake.[19] The components were then categorised into sex-specific quintiles and ranked from 1 to 5 if belonging to the ‘healthy group’ and from 5 to 1 otherwise. Since the intake of SSB was generally low (interquartile range, IQR: 1-to-7 servings/week) compared to the US population, where the score was developed, we used sex-specific tertiles of the distribution to rank this specific component from 3 to 1. Finally, the total DASH-score was calculated as the sum of all ranks, with the total score ranging from 8 to 36.

### Deriving RRR-scores

Prior to using the RRR method to derive DPs,[8] the 229 FFQ items were categorized into 32 groups based on between-food similarity (**Supplementary Table S2** ) and adjusted for sex-specific TEI using the nutrient residual method.[19] We then derived RRR-DPs separately for males and females to capture sex-specific dietary profiles associated with clinical and biochemical markers with known risk for CKD. Selected mediators, chosen for their known association with kidney health were: glycated haemoglobin (HbA1c), mean arterial pressure (MAP),[20] C-reactive protein (CRP), uric acid, total cholesterol (TC), ferritin, fibrinogen, serum potassium, and haemoglobin (HGB). Quantile normalization to the last assay was used to account for changes in measurement methods.[21, 22] To prevent confounding, all mediators were age-adjusted and, by means of multilevel models with random intercept for household, we controlled for family-related lifestyle or related environmental effects. The optimal number of DPs was identified via screeplot inspection, resulting in 2 DPs. Each DP represented a linear combination of the effects of the food groups on the mediators, summarized through a set of factor loadings. A dietary score (RRR-score) was calculated using the estimated factor loadings as weights for the standardized intake of each food item.[23] RRR modelling and optimal rank estimation were performed using the R package ‘rrpack’ v0.1-12.[24]

### Assessment of covariates

Potential confounders were identified via Directed Acyclic Graph (DAG) analysis (**Supplementary Figure S1**) using the online tool DAGitty (https://www.dagitty.net/dags).[25] We considered: age; sex; BMI (derived from weight and height measured at the study centre); physical activity (continuous Metabolic Equivalent of Task; MET);[26, 27] smoking habit (Current, Former, Never smoker);[28] education level (Primary school or no title; Lower secondary school; Vocational school; Upper secondary school; and University or higher); and special diet (Yes, No; indicating whether participants reported following a special diet for medical reasons). To account for non-independency of observations due to relatedness, models were adjusted for the first 10 principal components of the genotyped data with minor allele frequency >5%.[29] Since menstruation cessation was targeted as a potential effect modifier in the diet-eGFR association, females were classified in two groups based on their menstrual status: Menstruation_Yes_ and Menstruation_No_ according to their response to the question “*Do you still have regular menstrual bleedings?*”. Individuals with missing information on menstrual status (N=32) were assigned Menstruation_No_ if >50 years old and Menstruation_Yes_ otherwise.

### Statistical analyses

To describe and compare the DPs, we estimated the Pearson correlation coefficients between all the DP-scores, including the DASH-score, and between each age-adjusted RRR food group and the scores.

The relationship between the DASH-score, RRR-scores, and eGFR was analysed through linear regression models. To allow direct comparison of the estimates given the different scales of the scores, all dietary scores were z-standardized (mean=0, variance=1), allowing associations to be interpreted as per 1 standard deviation (1-SD) change. To reflect differences both in the DPs and disease propensity between males and females, only sex-specific analyses were performed. As informed by the DAG (**supplementary Figure S2** ), all models were adjusted for age, TEI, physical activity, smoking habits, education, BMI and special diet. To allow for potential non-linear effects, we additionally fitted generalized additive models (GAMs) with penalized regression splines for age and DP-scores, with smoothing parameter λ derived via restricted maximum likelihood, using the R package ‘mgcv’ v1.8-40.[30]

To investigate the potential influence of menstruation cessation on the relationship between diet and eGFR in females, we performed a preliminary analysis including an interaction term between DASH diet, DP-scores and menstrual status (Menstruation_Yes_; Menstruation_No_). To exclude the possibility of a modification effect of age, an interaction between DP-scores and age was also included in the models. Following the observed evidence that menstrual status was an effect modifier for one of the DP-scores (see **Supplementary Table S3** with dedicated information on the interaction analysis, and Results section), all analyses in females were additionally stratified accordingly.

To control for potential residual confounding driven by dietary changes due to any sub-clinical health conditions, we conducted a sensitivity analysis excluding participants with sub-clinical diabetes mellitus, reduced kidney function or microalbuminuria (HbA1c>6.5 mmol/mol or eGFR<60 mL/min/1.73m^2^ or urinary albumin-to-creatinine ratio (UACR)>30 mg/g), which we refer to as the Healthy+ sample.

All analyses were performed using the R software v4.1.1.[31]

## Results

### Participant characteristics

Our sample included 3278 females and 2855 males of similar age around 40 years old (**Table 1**). Females had lower median eGFR than males (99.8 versus 104.5 ml/min/1.73m^2^). Levels of selected mediator biomarkers were generally higher in males, except for CRP and fibrinogen, which were higher in females, and HbA1c and TC, which were equally distributed between sexes. When stratifying females by menstrual status, we observed higher levels of all biomarkers in females with ceased menstruation (Menstruation_No_) except for CRP. As expected, due to higher age (57.2 versus 32.8 years), this group had lower median eGFR (88.4 versus 104.1 ml/min/1.73m^2^; **Table 1**).

**Table 1:**
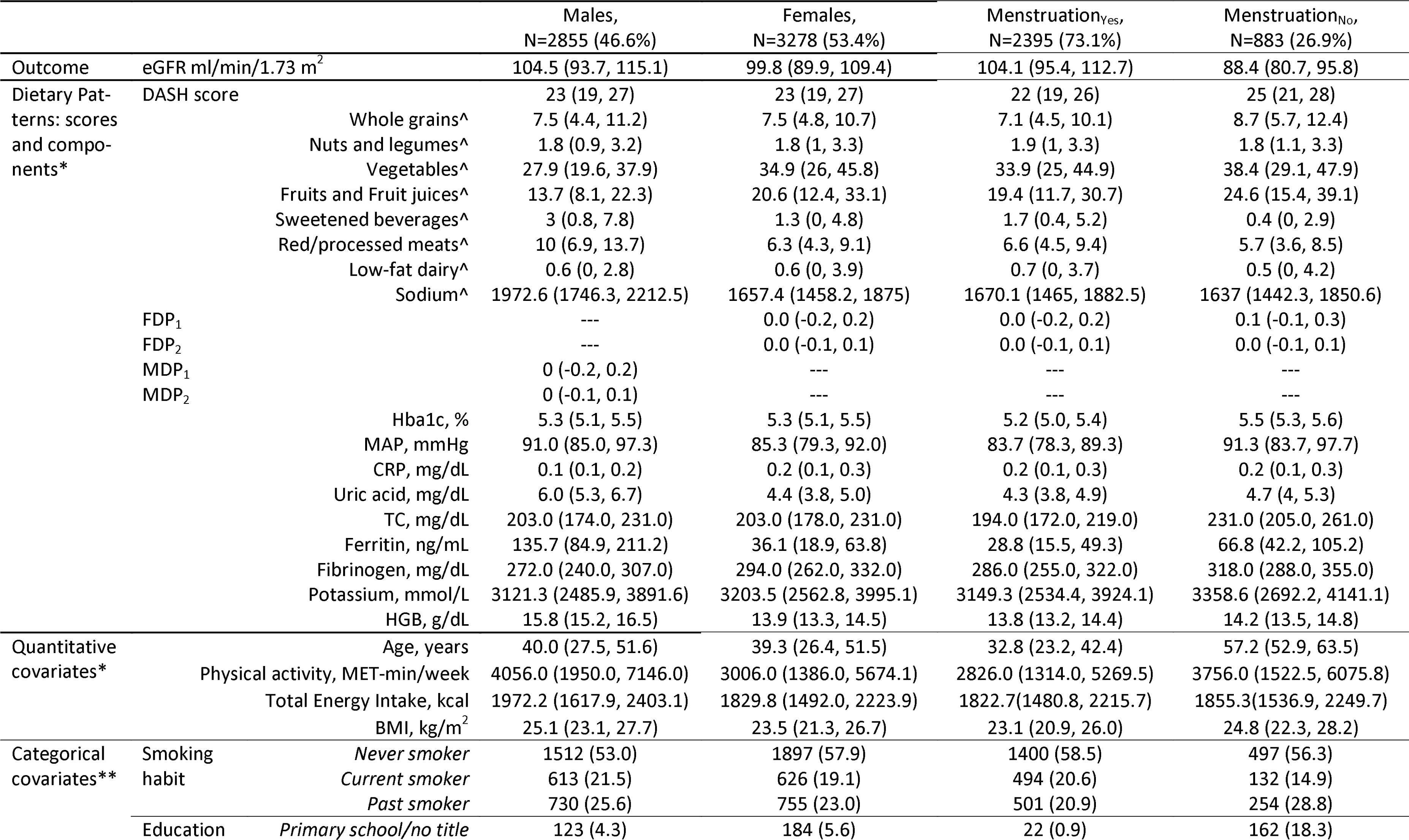
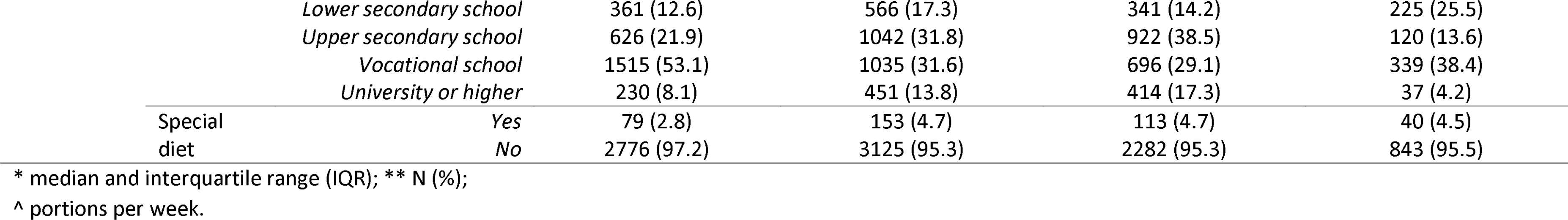
Sex-stratified characteristics of the study sample.

### Description of the DASH score, RRR-scores and their correlations

The overall DASH-score was equally distributed among sexes by design, as it was derived from sex-specific food intake quintiles. With respect to the DASH-score relevant dietary components, we observed a higher intake of ‘healthy’ food items (nuts and legumes, vegetables, fruit and fruit juices) in females, while males reported a higher intake of ‘unhealthy’ items, especially sodium, and red and processed meat (**Table 1**). Stratification of females by menstrual status revealed relevant differences in dietary habits, with lower intake of sodium, red and processed meat, and SSB after menstruation cessation, combined with higher intake of refined grains, vegetables, and fruits and fruit juices.

A general description of all dietary components used to derive RRR-scores is provided in **Supplementary Table S4**. We identified two RRR-scores for males and two different ones for females, which we labeled as MDP_1_ and MDP_2_ and FDP_1_ and FDP_2_, respectively (**Figure 1**). A high MDP_1_ reflected lower levels of all biomarkers. It was driven by higher intake of whole grains and cereals, sugar, fruits, and legumes, and lower intake of beer, and red and processed meat. High MDP_2_, which was associated with higher levels of HGB and HbA1c, and lower levels of TC, serum potassium, and ferritin, was mainly driven by low intake of coffee, alcohol (wine and beer), fish, fat, vegetable-based oils, and vegetables. In females, high FDP_1_ was associated with lower levels of uric acid, MAP, fibrinogen, ferritin, and CRP. FDP_1_ was driven by higher intake of whole grains, and dairy products, and by low intake of red and processed meat, refined grains, fish, poultry and alcohol (beer and spirits). High FDP_2_, which corresponded to higher levels of uric acid, HGB, and ferritin levels, and low levels of TC, MAP, HbA1c, fibrinogen and CRP, was driven by higher intake of beef, nuts, beer, legumes, fish, and coffee, and lower intake of refined grain.

**Figure 1:**
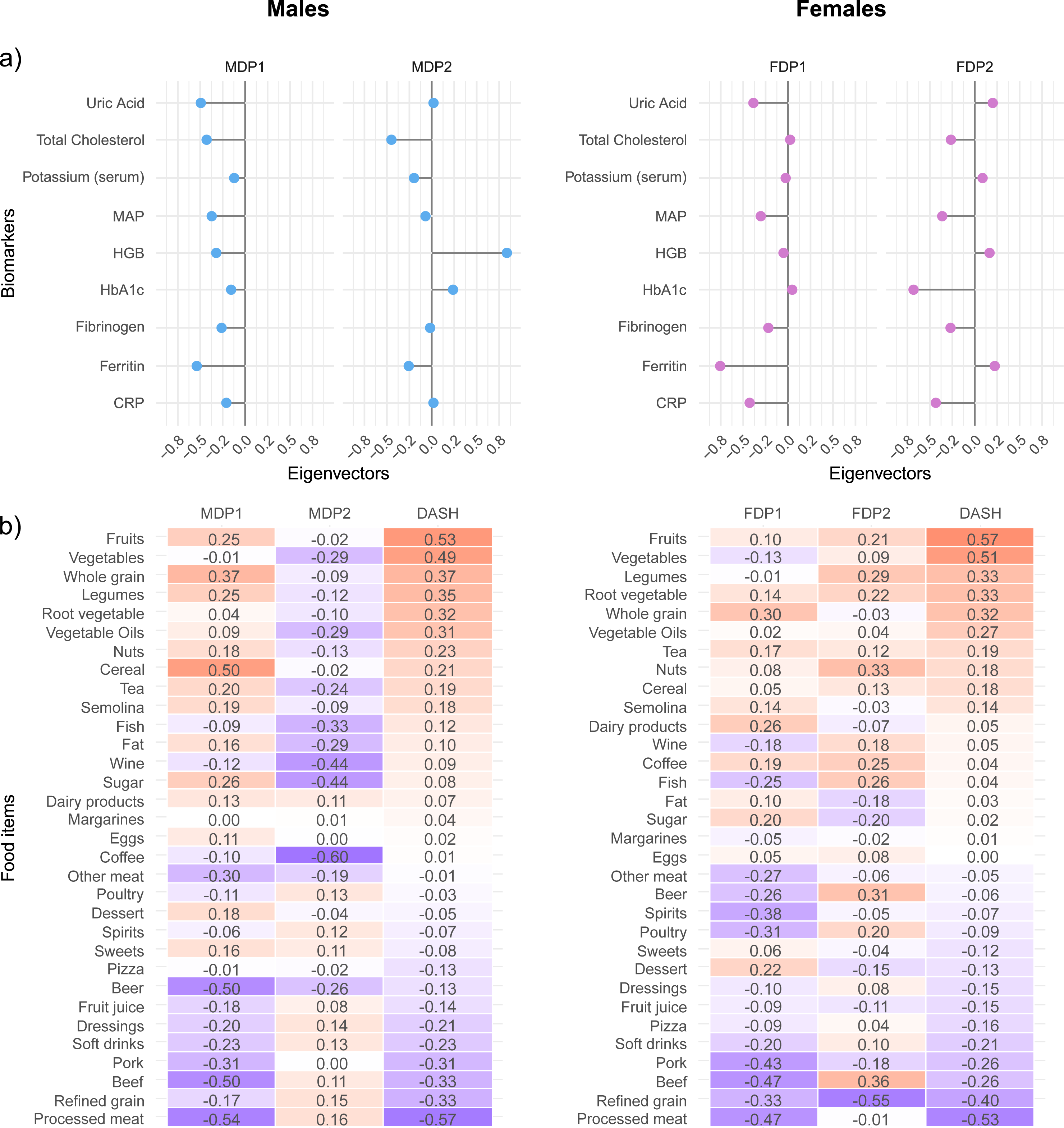
Representation of the Dietary Patterns (DPs). a) RRR-based eigenvectors summarizing the influence of the biomarkers in defining the respective DP-scores in males and females. b) Heatmaps of the Pearson’s correlation coefficients between RRR food groups and DP scores.

While the correlations between RRR-scores were null by design, in males, we observed moderate positive correlations of the DASH-score with MDP_1_ (r_DASH-MDP1_=0.52) and a weak negative correlation with MDP_2_ (r_DASH-MDP2_=-0.27). In females, we observed a moderate positive correlation of the DASH-score with FDP_1_ (r_DASH-FDP1_=0.34) and a weak positive correlation with FDP_2_ (r_DASH-FDP2_=0.18).

### Associations between DASH-score, RRR-scores and eGFR

The preliminary analysis in females(**Supplementary Table S3** ) investigating the potential modification effects of age and menstrual status, showed a significant interaction between menstrual status and FDP_1_ (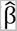: 2.12 ml/min/1.73 m^2^; 95%CI: 0.85, 3.40).

Final associations between DPs and eGFR are summarized in **Table 2**. In males, each 1-SD increase in the DASH-score was associated with 0.64 ml/min/1.73 m^2^ (estimated association coefficient, 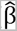) higher eGFR (95%CI: 0.22, 1.06). The GAM’s estimated degrees of freedom (edf) were 1.8, suggesting a slightly quadratic association (**Figure 2**). In females, no association was found between the DASH-score and eGFR, independent of menstrual status.

**Figure 2:**
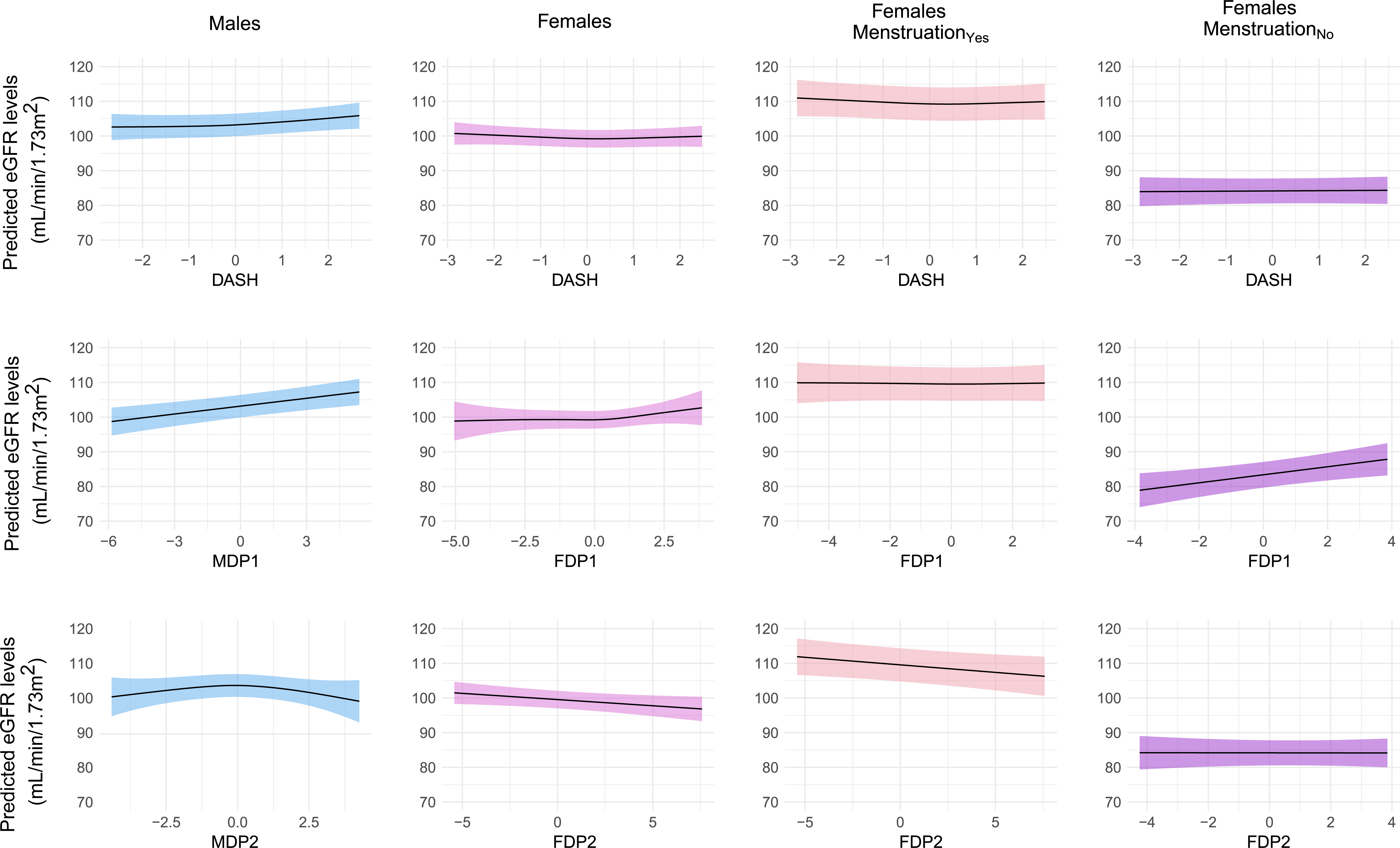
Plots of predicted eGFR levels by the different scores, as obtained from the fitted GAMs.

**Table 2:**
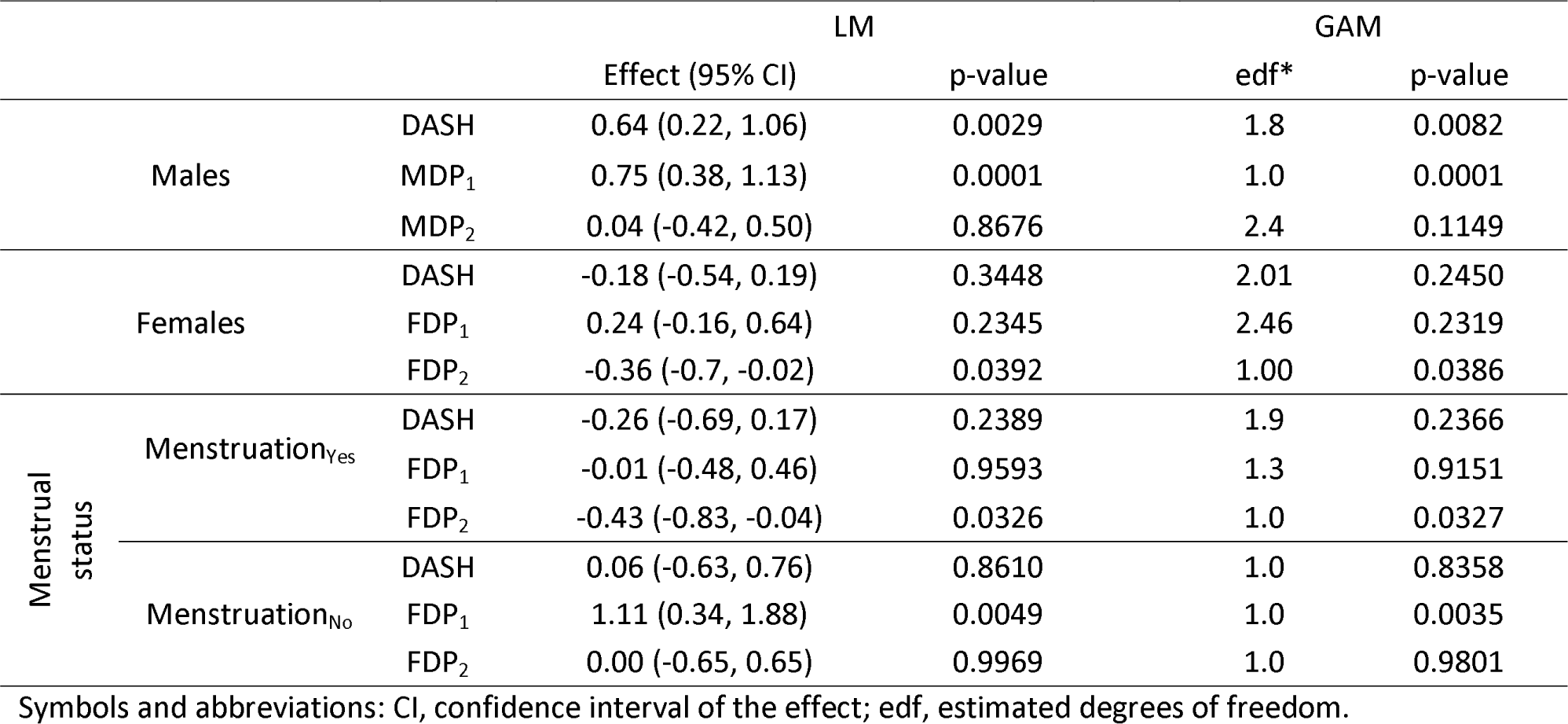
Results of the linear (LM) and generalized additive models (GAM). LM: Effects of DPs on eGFR in ml/min/1.73 m with 95% confidence intervals (CI) and p-values; GAM: estimated degrees of freedom (edf) and p-values.

Regarding the RRR-scores in males, MDP_1_ was positively associated with eGFR (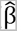=0.75 ml/min/1.73 m^2^; 95%CI: 0.38, 1.13); no association was observed between MDP_2_ and eGFR. In females, we observed negative associations between FDP_2_ and eGFR, both overall (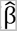: -0.36 ml/min/1.73 m^2^; 95%CI: -0.70, -0.02), and in the Menstruation group (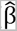: -0.43 ml/min/1.73 m^2^; 95%CI: -0.83, -0.04). In the Menstruation_No_ group, results showed a positive association between FDP_1_ and eGFR (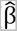: 1.11 ml/min/1.73 m^2^; 95%CI: 0.34, 1.88). The GAMs confirmed both significance and linearity of the results, with 1 edf for all significant associations (**Figure 2**).

### Sensitivity analyses

The Healthy+ subgroup did not differ substantially from the overall sample (**Supplementary Table S5** ). Results between the DPs and eGFR remained stable in the Healthy+ subgroups (**Supplementary Table S6**). In males, we confirmed the positive association with eGFR for the DASH-score (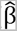: 0.67 ml/min/1.73 m^2^; 95%CI: 0.24, 1.09) and MDP (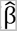: 0.83 ml/min/1.73 m^2^; 95%CI: 0.44, 1.21), whereas again no association was seen between MDP_2_ and eGFR. In females, FDP_2_ was negatively associated with eGFR both overall (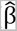: -0.39 ml/min/1.73 m^2^; 95%CI: -0.73, -0.04), and in the Menstruation group (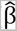: -0.46 ml/min/1.73 m^2^; 95%CI: -0.87, -0.05). In the Menstruation group, FDP appeared to be linearly associated with eGFR in the GAM, however this association was not confirmed by the linear model. In all models we observed a slight increase in the magnitude of the significant associations. GAMs confirmed both linearity and significance of those associations.

## Discussion

In this investigation, we applied two different methodological approaches to estimate DPs, providing novel insights into the relationship between diet and kidney function in the general population. Our main findings suggest a role of sex and menstrual status in CKD prevention through diet. In males, a diet with higher intake of cereals, whole grains, sugar, fruits, and legumes, and low intake of beer, red and processed meat, was associated with higher eGFR, reflecting better kidney function. A similar association was observed for high adherence to a DASH-style diet. In females, the associations between diet and eGFR varied by menstrual status. After menstruation cessation (Menstruation_No_) a DP characterized by low intake of any kind of meat, alcohol (beer and spirits) and fish, and high intake of whole grains and dairy products was associated with a higher eGFR. Before menstruation cessation (Menstruation_Yes_) higher intake of beef, nuts, beer, legumes, fish, and coffee, and a low intake of refined grains was found to be associated with lower eGFR. This latter result was consistent with the observations in the overall group of women.

The DASH diet has been favourably linked to relevant risk factors associated with CKD onset, such as insulin resistance, hypertension, and dyslipidemia, and might therefore also be protective against kidney dysfunction.[4, 18] A recent meta-analysis of six observational studies estimated that higher adherence to the DASH-score was associated with a 23% lower risk of developing CKD.[5] However, a negative association between DASH-style diet and CKD risk was observed only in studies where the index was extracted from nutrients, but not food groups. In contrast to these, we found that higher adherence to the food group-based DASH-score was associated with higher eGFR in males only, reflecting better kidney function. Although the DASH-score was derived from sex-specific quintiles, hence similarly distributed in males and females, we observed large differences in the actual consumption of the specific DASH dietary components. Specifically, higher consumption of ‘healthy’ food items (nuts and legumes, vegetables, fruit and fruit juices) was observed in females, and higher consumption of ‘unhealthy’ items, especially sodium, and red and processed meat in males. This could to some extent explain the sex-related discrepancies in our findings.

To the best of our knowledge, this is the first investigation using CVD and CKD risk biomarkers to identify disease-oriented DPs using RRR in a generally healthy population. Other studies applying RRR have selected different sets of biomarkers, as well as different kidney outcomes. In a population-based cohort in the north of the Netherlands, Cai and colleagues[32] derived RRR-based DPs directly related to eGFR at baseline and evaluated its association with eGFR decline and CKD incidence at five-year follow-up. Among females, they observed a DP characterized by high consumption of eggs, cheese, and legumes, and low consumption of sweets, white meat, and commercially prepared dishes. In males, the DP was characterized by high consumption of cheese, bread, milk, fruits, vegetables, and beer and low consumption of white and red meat. Higher adherence to the scores was associated with lower risk of eGFR decline in both sexes, which aligns with our results. Using health screening data from a health institute in Taiwan, Kurniawan et al.[10] estimated kidney function-oriented DPs including waist-to-hip ratio, triglycerides, LDL-C, TC/HDL-C, calcium, phosphorus, creatinine, and blood urea nitrogen as mediators, and identified a corresponding DP associated with increased risk of impaired kidney function among older individuals with CKD. High adherence to this DP was characterized by higher intake of preserved vegetables, processed meat or fish, rice or flour products, meat, soy sauce, organ meats, fried rice or flour products, and instant noodles, along with lower intakes of fruits, dark-coloured vegetables, bread, and beans or bean products. Although we used a different set of biomarkers and conducted the analyses among a population free of CKD and related comorbidities, our results are at least partially concordant with their findings regarding the obtained DPs and the association with kidney function, specifically in males.

Overall, in both sexes any kind of meat and fish, as well as whole grains, fruits, vegetables, legumes, and nuts, played a strong role in the RRR-based DP composition. Animal-sourced proteins, processed foods, and refined grains are known to result in high acidic loads when metabolized,[10] in contrast with vegetable-sourced proteins, which lead lower acidic loads.[33] High dietary acidic load may consequently induce kidney adaptive mechanisms to increase acid excretion, thereby promoting renal injury.[34] Moreover, a higher plant-to-animal protein ratio in the diet may protect against kidney dysfunction by increasing serum bicarbonate levels and reducing fibroblast growth factor 23 levels.[5, 35] We have observed a similar impact of poultry and fish consumption in two of our previous analyses investigating associations between nutrient-related DPs and kidney function and damage[36] as well as the impact of different protein sources on eGFR.[37] Although regular fish intake is suggested to promote general and specifically cardiovascular health, some uncertainties remain regarding its health benefits, which could be related to the type and preparation of the fish and whether it is processed.[38] Moreover, fruit and vegetable consumption may exert beneficial effects on kidney function through higher intake of fiber, magnesium and potassium. Low fiber consumption has been associated with increased inflammatory cytokines which may contribute to impaired kidney function.[5] Nuts and legumes are rich in magnesium and potassium. Higher magnesium intake might preserve kidney function through reduction of endothelial disfunction ,[39] while higher potassium intake could contribute through the mitigation of blood pressure levels.[40]

In females, we found potential evidence of the diet-eGFR association to be modified by menstrual status. Specifically, we observed a negative association between FDP_2_ and eGFR in females who reported still having regular menstrual bleedings. In females with ceased menstruation, we observed a positive association between FDP_1_ and eGFR. These differences might suggest a role of iron metabolism and ferritin levels that could modify the relationship between diet and kidney function in females. A recent Japanese study revealed that both low and high serum ferritin levels are linked to increased risk of adverse kidney outcomes compared to intermediate levels.[41] Elevated serum ferritin levels, in particular, may pose a risk of iron overload.[42, 43] Moreover, ferritin is closely related to other inflammation markers.[44] Together with ferritin, in fact, CRP and fibrinogen were also relevant for FDP_1_ profile, supporting evidence from previous studies suggesting a potential role of inflammation on kidney function levels.[45] In addition, the concurrent presence of high serum uric acid and CRP levels could be associated with an increased risk of type II diabetes.[46] Therefore it seems reasonable to observe higher eGFR levels in individuals with a dietary profile (FDP_1_) associated with lower levels of these markers. Nevertheless, when interpreting these results, it is important to remember that baseline levels of biomarkers differed by menstrual status, with higher levels of all biomarkers observed in females with ceased menstruation, except for CRP.

The comparison of the two approaches to define the DPs revealed a positive correlation between the RRR-based MDP_1_ and the DASH score. This suggests that our population-specific, disease-oriented DPs, based on several markers related to CVD, do reflect, to some extent, the dietary guidelines defined to contrast hypertension (DASH) in males. Overall, the RRR-scores were more strongly associated with eGFR than the DASH-score. This might be due to the flexibility of the approach, allowing to shape DPs in a disease-related manner, potentially leading to the detection of unknown associations and complex mechanisms. For example, FDP_2_ was weakly correlated with the DASH-score, but it highlighted a relevant pattern related to lower eGFR in the Menstruation_Yes_ group. Despite being beyond the scope of our investigation, it is worth considering the appropriateness of selecting common biomarkers for males and females, particularly given recent studies highlighting sex differences in disease biomarkers, as observed in CVDs.[47] This further emphasizes the recent recognition of sex differences in the aetiology, mechanisms, epidemiology, and treatment of CKD.[48] Moreover, females are less likely to receive diagnoses, monitoring, and management compared to males.[49] While evidence suggests that sex influences various aspects of treatment and complications in later-stage kidney disease, research on its impact on earlier CKD stages is limited.[50, 51] While CKD is more prevalent in females,[13] kidney function declines more rapidly in males.[52] It has been hypothesized that these sex differences may be attributed to the protective effects of endogenous estrogens against the deleterious effects of testosterone on kidney function and structure, as well as the generally healthier lifestyles observed in females.[48] In addition, the role of menstrual status, probably mediated by ferritin and inflammation markers, underscores the need for further investigation concerning CKD in females.

The main strength of our investigation is the large sample size, which enabled us to conduct sex-stratified analyses without sacrificing statistical power. Next, diet was assessed through a validated FFQ. Third, we were able to include a wide set of parameters, including measured biochemical markers and information collected through self-reporting or by trained study-nurses. In particular, detailed information on medical history and lifestyle allowed us to control for an extensive range of relevant confounders. Finally, we included and compared two approaches to evaluate DPs in our sample. This enabled us to obtain a complementary and comprehensive overview of the role of diet in the association with eGFR. While using the DASH index simplified the interpretation and comparison of our findings with other studies, the complementary usage of a hybrid approach as RRR allowed as to derive our DPs in a more flexible and exploratory way. Specifically, it allowed us to consider the multifactorial and complex nature of CKD, often characterized by multiple comorbidities, which could share aspects of their pathophysiology and require the formulation of comprehensive dietary recommendations.

The main limitation of our investigation is the cross-sectional nature of the data. In fact, the simultaneous collection of health information and dietary intake could prevent causal interpretations of our findings. However, our choice of focusing on individuals who self-reported being free from CKD, hypertension, or diabetes, along with adjustments made for those following a special diet, may mitigate the risk of reverse causation. Moreover, the robustness of our results following the exclusion of individuals with subclinical diabetes mellitus (Healthy+), reduced kidney function, or microalbuminuria, confirms that, given the silent nature of CKD, especially in its early stages, it is unlikely to stimulate lifestyle changes prior to formal diagnosis. Another limitation is the absence of measured GFR and cystatin C in our study: GFR was estimated based on serum creatinine, which is also influenced by muscle mass and protein intake. This could have affected our results, especially those related to the consumption of animal-based foods, which are rich in protein. However, the use of 12-hour fasting plasma samples, as done by the CHRIS study, has been shown to mitigate the acute effect of meat consumption on serum creatinine levels.[53] Other limitations are related to the use of an FFQ to assess dietary intake. The first is represented by the common challenge of extrapolating sodium intakes.[54] This could have led to an underestimation of detrimental effects of lower adherence to the DASH diet, as well as other processed food items, rich in sodium and other additives.[55] Moreover, dietary intake was assessed only once, referring to the average intake of participants potentially leading to recall bias.

## Conclusion

A DASH-style diet and a disease-oriented dietary pattern characterized by higher intake of whole grains, cereals, fruits and legumes, and lower intake of beer, red and processed meat, SSBs and fruit juice, were associated with higher eGFR in males without CKD, diabetes and hypertension. In females, associations between RRR-based DPs and eGFR were modified by menstrual status. Overall, results highlight the need to consider sex-differences when studying the relationship between diet and kidney health.

## Data Availability

Data and samples can be requested for clearly defined research via the CHRIS Portal (CHRIS Portal - Eurac Research: https://chrisportal.eurac.edu/).

## Acknowledgements

CHRIS Study investigators thank all study participants, the Healthcare System of the Autonomous Province of Bolzano-South Tyrol, and all Eurac Research staff involved in the study (CHRIS acknowledgements

<u>Eurac Research</u>) Bioresource Impact Factor Code: BRIF6107. The authors thank the Department of Innovation, Research University and Museums of the Autonomous Province of Bozen/Bolzano for covering the Open Access publication costs.

## Data availability statement

Data and samples can be requested for clearly defined research via the CHRIS Portal (CHRIS Portal -

Eurac Research).

## Ethics approval and consent to participate

The Ethics Committee of the Healthcare System of the Autonomous Province of Bolzano-South Tyrol approved the CHRIS baseline protocol on 19 April 2011 (21-2011). The study conforms to the Declaration of Helsinki, and with national and institutional legal and ethical requirements. All participants included in the analysis gave written informed consent.

## CRediT author statement

**Giulia Barbieri**: Conceptualization, Investigation, Data curation, Formal analysis, Methodology, Visualization, Writing Original Draft; **Vanessa Garcia-Larsen**: Conceptualization, Writing Original Draft; **Rebecca Lundin**: Methodology, Writing Original Draft; **Ryosuke Fujii**: Methodology, Writing Original Draft; **Pietro Manuel Ferraro**: Methodology, Writing review & editing; **Giovanni Gambaro**: Methodology, Writing review & editing; **Roberto Melotti**: Methodology, Writing review & editing; **Martin Gögele**: Data curation, Writing review & editing; **Lucia Cazzoletti**: Conceptualization, Writing review & editing

**Peter P Pramstaller**: Funding acquisition, Writing review & editing; **Maria Elisabetta Zanolin**: Funding acquisition, Conceptualization, Writing review & editing; **Cristian Pattaro**: Conceptualization, Funding acquisition, Supervision, Writing Original Draft; **Essi Hantikainen**: Conceptualization, Methodology, Investigation, Supervision, Writing Original Draft

