## Supplementary material for "DASH diet, Reduced Rank Regression Dietary Patterns and relations with kidney function in the CHRIS general population study"

### **Table S1**: ATC codes used to identify self-reported medications for hypertension and diabetes

| Type of medication | Therapy | ATC code |
| --- | --- | --- |
| Blood pressure lowering drugs | Antihypertensives | C02AC01, C02CA04, C02LA01 |
|  | Diuretics | C03AA03, C03BA08, C03BA11, C03CA01, C03CA04, C03DA01, C03DA02, C03DA04, C03EA01, C03EB01 |
|  | Peripheral vasodilators | C04AD03, C04AE01, C04AE51 |
|  | Beta blocking agents | C07, C07AA05, C07AA06, C07AA07, C07AB02, C07AB03, C07AB07, C07AB12, C07AG02, C07B, C07BB12, C07CB03 |
|  | Calcium channel blockers | C08CA01, C08CA02, C08CA05, C08CA09, C08CA11, C08CA12, C08CA13, C08DA01, C08DB01 |
|  | Agents acting on the renin-angiotensin system | C09AA02, C09AA03, C09AA04, C09AA05, C09AA06, C09AA07, C09AA09, C09AA15, C09BA01, C09BA02, C09BA03, C09BA04, C09BA05, C09BA06, C09BA09, C09BA12, C09BA15, C09BB02, C09BB04, C09BB07, C09BX01, C09CA01, C09CA03, C09CA04, C09CA06, C09CA07, C09CA08, C09DA, C09DA01, C09DA03, C09DA04, C09DA06, C09DA07, C09DA08, C09DB02 |
| Drugs used in diabetes | Insulins and analogues | A10AB04, A10AB05, A10AB06, A10AD05, A10AE04, A10AE05, A10AE06 |
|  | Blood glucose lowering drugs (excl. insulins) | A10BA02, A10BB09, A10BB12, A10BD02, A10BD05, A10BD06, A10BD07, A10BD08, A10BD13, A10BG03, A10BH01, A10BH03, A10BK01, A10BX02, A10BX07 |

### **Table S2**: Composition of the food groups

|  | **Specific food** |
| --- | --- |
| **RRR food groups** | |
| Beef | Braising steak, Burger, Mince, Stewing steak, Veal escalope |
| Beer | Beer |
| Cereal | Breakfast cereal |
| Coffee | Coffee |
| Dairy products | Cow milk (fresh and sour), Sheep or goat milk, Lactose-free milk, Yogurt (whole milk and Greek), Kefir, Cheese, Cream (fresh and sour) |
| Dessert | Cakes, Pastries, Muffins, Doughnuts, Pudding, Homemade Biscuits, Ice cream |
| Dressings | Salad dressing, Mustard, Mayonnaise, White, Tomato ketchup |
| Eggs | Omelette, Eggs |
| Fat | Normal butter (70% fat), Lard |
| Fish | Tuna, Crab, Salmon, Cod, Roe, Mussels, Breaded fish fillet |
| Fruit juice | Fruit juice drink/squash (with and without sugar) |
| Fruits | Capers, Apples, Pears, Bananas, Avocado, Cherries, Rhubarb, Blueberries, Melon, Grapes, Mangoes, Apricots, Peaches, Nectarines, Plums, Prunes, Orange juice, Pineapple, Kiwi fruit, Lemon juice, Oranges, Mandarin oranges, Grapefruit, Raisins, Figs, Olives, Kaki |
| Legumes | Beans, Lentils, Beans, Green peas, Soy milk, rice milk, oat milk, Tofu |
| Margarines | Normal margarine, Blended spreads, Soya-based margarine or spreads |
| Nuts | Peanuts, Cashew nuts |
| Other meat | Rabbit, Lamb, Pate, Tongue |
| Pizza | Pizza |
| Pork | Pork (smoked, chop, steak, fillet, rips) |
| Poultry | Chicken,  Turkey |
| Processed meat | Sausages, Salami, smoked ham, Vienna sausage, Bacon, Any smoked/cured poultry |
| Root vegetable | Carrots, Parsnip, Turnip, Beetroot, Celery, Potatoes, Fried potato, Gnocchi, Potato cakes, Sweet potato |
| Semolina | Couscous in savoury dishes, Polenta |
| Soft drinks | Carbonated lemonade, Normal carbonated/soft/isotonic drinks, Diet carbonated/soft/isotonic drinks, Energy drinks |
| Spirits | Sherry, Spirits |
| Sugar | Sugar, Jam, Honey |
| Sweets | Wholemeal biscuits, sweet/spicy biscuits (e.g. cream, jam, chocolate), Sweets, Cereal bars, Lollies, Chocolate bars, Chocolate (milk), Spreads with nuts |
| Tea | Tea (black, green and herbal) |
| Vegetable Oils | Sunflower, Olive, Extra virgin olive, Rape seed, Lin seed, Pumkin seed |
| Vegetables | Lettuce, Spinach, Chard, Amaranth leaves, Okra, Tomatoes, Aubergine, Courgette, Peppers, Capsicum, Cucumber, Artichoke, Radish leaves, Coleslaw, Sweetcorn, Asparagus, Mixed herbs, Leeks, Mushrooms, Onions, Garlic, Cauliflower, Pumpkin, Brussels sprouts, Broccoli, Cabbage, Gherkins, Ginger |
| White grain | Bread (white and naan), Chapatis, Pasta, Noodles, White rice, Noodles |
| Whole grain | Bread (brown), Rye crispbread, Rusks, Crackers, Pasta (wholewheat), Brown rice |
| Wine | Wine |
| **DASH food groups** | |
| Fruits and Fruit juices | Capers, Apples, Pears, Bananas, Avocado, Cherries, Rhubarb, Blueberries, Melon, Grapes, Mangoes, Apricots, Peaches, Nectarines, Plums, Prunes, Orange juice, Pineapple, Kiwi fruit, Lemon juice, Oranges, Mandarin oranges, Grapefruit, Raisins, Figs, Olives, Kaki, fruit juice/smash without sugar |
| Low-fat dairy | Low-fat milk, Skimmed milk, Lactose-free milk, Kefir, Cottage cheese |
| Nuts and legumes | Peanuts, Cashew nuts, Beans, Lentils, Beans, Green peas, Soy milk, rice milk, oat milk, Tofu |
| Vegetables | Lettuce, Spinach, Chard, Amaranth leaves, Okra, Tomatoes, Aubergine, Courgette, Peppers, Capsicum, Cucumber, Artichoke, Radish leaves, Coleslaw, Sweetcorn, Asparagus, Mixed herbs, Leeks, Mushrooms, Onions, Garlic, Cauliflower, Pumpkin, Brussels sprouts, Broccoli, Cabbage, Gherkins, Ginger, Carrots, Parsnip, Turnip, Beetroot, Celery, Potatoes, Fried potato, Gnocchi, Potato cakes, Sweet potato |
| Red and processed meats | Sausages, Salami, smoked ham, Vienna sausage, Bacon, Any smoked/cured poultry, Braising steak, Burger, Mince, Stewing steak, Veal escalope, Pork (smoked, chop, steak, fillet, rips), Liver pate, Other offal (tongue, brain, heart, kidney, tripe, stomach), Rabbit, Pheasant, Duck |
| Sweetened beverages | Concentrated fruit juice, with sugar, Carbonated lemonade, Normal carbonated/soft/isotonic drinks, Diet carbonated/soft/isotonic drinks, Energy drinks |
| Whole grains | Bread (brown), Rye crispbread, Rusks, Crackers, Pasta (wholewheat), Brown rice |
| Sodium | Total daily Sodium (mg) |

### **Table S3**: Model to assess the potential mediating role of menstruation status in the diet-eGFR relationship*. Main and interaction effects, 95% confidence intervals (95%CI) and p-value.

|  | Main effect | | Interaction with the Dietary Pattern (DP) | |
| --- | --- | --- | --- | --- |
|  | Effect (95% CI) | P-value | Effect (95% CI) | P-value |
| *DASH* | 0.53 (-0.64, 1.70) | 0.3740 |  |  |
| *Menstruation Status* | -0.18 (-1.33, 0.97) | 0.7558 | 0.81 (-0.32, 1.95) | 0.1603 |
| *Age* | -0.66 (-0.7, -0.62) | 6.45E-235 | -0.02 (-0.06, 0.01) | 0.1671 |
| *FDP_1_* | 1.07 (-0.16, 2.31) | 0.0879 |  |  |
| *Menstruation Status* | -0.46 (-1.62, 0.70) | 0.4349 | 2.12 (0.85, 3.40) | 0.0011 |
| *Age* | -0.66 (-0.7, -0.63) | 1.70E-230 | -0.03 (-0.07, 0.00) | 0.0520 |
| *FDP_2_* | -0.81 (-1.93, 0.30) | 0.1522 |  |  |
| *Menstruation Status* | -0.13 (-1.26, 1.00) | 0.8209 | 0.05 (-1.02, 1.12) | 0.9240 |
| *Age* | -0.66 (-0.7, -0.62) | 8.46E-240 | 0.01 (-0.02, 0.04) | 0.4822 |

*Underlying model: $eGFR=DPscore+Age+Menstruation status+ DPscore:Age+ DPscore:Menstruation status+other covariates$

### **Table S4**: Median and interquartile range of food intakes (portions per weeks) for the RRR-based food groups

| Food intake | Males,  N=2855 (46.6%) | Females,  N=3278 (53.4%) | Menstruation_Yes_,  N=2395 (73.1%) | Menstruation_No_,  N=883 (26.9%) |
| --- | --- | --- | --- | --- |
| Whole grains | 7.53 (4.4, 11.17) | 7.5 (4.8, 10.7) | 8.7 (5.7, 12.4) | 7.1 (4.5, 10.1) |
| Refined grains | 8 (5.55, 10.99) | 5.7 (3.7, 8.3) | 5 (3.2, 7.7) | 6 (4, 8.4) |
| Cereal | 0 (0, 0.88) | 0.4 (0, 1.6) | 0 (0, 0.8) | 0.6 (0, 1.8) |
| Semolina | 0.55 (0.15, 0.93) | 0.7 (0.3, 1.1) | 0.8 (0.4, 1.2) | 0.6 (0.3, 1) |
| Dessert | 3.59 (2.21, 5.51) | 3 (1.9, 4.8) | 2.7 (1.6, 4.5) | 3.1 (2, 4.8) |
| Eggs | 1.54 (1.02, 2.58) | 1.5 (0.9, 2.6) | 1.3 (0.8, 2.2) | 1.6 (1, 2.7) |
| Sweets | 3.82 (1.86, 7.21) | 3.9 (1.8, 7.1) | 2.5 (1.1, 5.2) | 4.3 (2.3, 7.7) |
| Sugar | 7.65 (2.9, 12.85) | 6.7 (2.8, 11.6) | 8.3 (4.7, 13.9) | 6.1 (2.1, 10.7) |
| Vegetable Oils | 8.88 (5.64, 13.19) | 10.6 (7.2, 15.2) | 12.1 (8.4, 16.9) | 9.9 (6.8, 14.5) |
| Margarines | 0 (0, 0) | 0 (0, 0) | 0 (0, 0) | 0 (0, 0.4) |
| Fat | 3.32 (0.88, 6.63) | 3.8 (1.1, 6.9) | 4.8 (2.1, 7.7) | 3.5 (1, 6.5) |
| Nuts | 0.28 (0, 0.78) | 0.3 (0, 0.8) | 0 (0, 0.6) | 0.3 (0, 0.8) |
| Legumes | 1.28 (0.59, 2.33) | 1.3 (0.7, 2.4) | 1.4 (0.8, 2.3) | 1.3 (0.7, 2.4) |
| Vegetables | 25.02 (17.59, 33.82) | 31.4 (23.2, 40.9) | 34.3 (26, 43) | 30.5 (22.5, 40.2) |
| Fruits | 12.85 (7.6, 21.43) | 19.7 (11.9, 32.3) | 24 (14.9, 38.6) | 18.5 (11.2, 29.8) |
| Root vegetable | 5.86 (4.26, 7.95) | 6.2 (4.6, 8.5) | 6.6 (5, 8.9) | 6.1 (4.5, 8.3) |
| Fruit juice | 0.98 (0, 3.93) | 0.7 (0, 3.1) | 0.2 (0, 2.1) | 0.9 (0, 3.4) |
| Soft drinks | 1.31 (0.24, 4.02) | 0.4 (0, 1.6) | 0 (0, 0.6) | 0.7 (0, 2) |
| Tea | 0.52 (0, 2.29) | 1.6 (0.4, 5.5) | 2.4 (0.5, 6.4) | 1.3 (0.4, 5.1) |
| Coffee | 8.06 (4.74, 12.5) | 7.5 (2.9, 12) | 8.5 (5.4, 12.5) | 6.9 (1, 11.8) |
| Beer | 1.09 (0.5, 2.89) | 0 (0, 0.6) | 0 (0, 0.5) | 0 (0, 0.6) |
| Wine | 0.93 (0, 2.31) | 0.6 (0, 1.3) | 0.6 (0, 2.3) | 0.5 (0, 1.1) |
| Spirits | 0 (0, 0.56) | 0 (0, 0.3) | 0 (0, 0) | 0 (0, 0.4) |
| Beef | 3.2 (2.14, 4.62) | 2.3 (1.6, 3.3) | 2.1 (1.4, 3) | 2.4 (1.6, 3.4) |
| Pork | 0.86 (0.47, 1.56) | 0.5 (0, 0.9) | 0.5 (0, 0.9) | 0.5 (0, 0.9) |
| Processed meat | 4.88 (2.75, 7.67) | 2.9 (1.6, 5) | 2.4 (1.2, 4.3) | 3 (1.7, 5.2) |
| Other meat | 0.15 (0, 0.65) | 0 (0, 0.4) | 0 (0, 0.5) | 0 (0, 0.3) |
| Poultry | 1.02 (0.57, 1.62) | 0.9 (0.5, 1.5) | 0.8 (0.4, 1.3) | 1 (0.5, 1.6) |
| Fish | 1.73 (0.94, 2.76) | 1.6 (0.9, 2.6) | 1.4 (0.8, 2.4) | 1.7 (0.9, 2.7) |
| Dairy products | 14.95 (10.47, 19.83) | 16.5 (12, 21.5) | 16.8 (12.3, 21.6) | 16.4 (11.9, 21.5) |
| Dressings | 2.11 (1.16, 3.68) | 1.6 (0.9, 2.7) | 1.3 (0.6, 2.3) | 1.7 (1, 2.9) |
| Pizza | 0.63 (0.48, 0.93) | 0.6 (0.4, 0.8) | 0.5 (0.4, 0.7) | 0.6 (0.5, 0.9) |

|  |  | Males, N=2775 (47.4%) | Females, N=3075 (52.6%) |
| --- | --- | --- | --- |
| Outcome  Dietary Patterns: scores and components* | eGFR, ml/min/1.73 m^2^ | 104.7 (94.1, 115.2) | 99.7 (89.9, 109.3) |
|  | DASH | 23.0 (19.0, 27.0) | 23.0 (19.0, 27.0) |
|  | Whole grains^ | 7.5 (4.4, 11.1) | 7.5 (4.8, 10.7) |
|  | Nuts and legumes^ | 1.8 (0.9, 3.2) | 1.9 (1.0, 3.3) |
|  | Vegetables^ | 28.0 (19.7, 37.9) | 34.8 (26.0, 45.9) |
|  | Fruits and Fruit juices^ | 13.7 (8.1, 22.4) | 20.6 (12.3, 33.2) |
|  | Sweetened beverages^ | 3.0 (0.8, 7.8) | 1.3 (0.0, 4.8) |
|  | Red and processed meats^ | 10.1 (7.0, 13.7) | 6.3 (4.2, 9.0) |
|  | Low-fat dairy^ | 0.6 (0.0, 2.9) | 0.6 (0.0, 4.1) |
|  | Sodium^ | 1972.6 (1746.3, 2211.7) | 1655.1 (1458.4, 1867.8) |
|  | FDP1 | --- | 0.0 (-0.2, 0.2) |
|  | FDP2 | --- | 0.0 (-0.1, 0.1) |
|  | MDP1 | 0.0 (-0.2, 0.2) | --- |
|  | MDP2 | 0.0 (-0.1, 0.1) | --- |
|  | Hba1c, % | 5.3 (5.1, 5.5) | 5.3 (5.1, 5.5) |
|  | MAP, mmHg | 91.0 (85.0, 97.0) | 85.0 (79.3, 92.0) |
|  | CRP, mg/dL | 0.1 (0.1, 0.2) | 0.2 (0.1, 0.3) |
|  | Uric acid, mg/dL | 5.9 (5.2, 6.7) | 4.4 (3.8, 5.0) |
|  | TC, mg/dL | 203.0 (174.0, 231.0) | 203.0 (178.0, 232.0) |
|  | Ferritin, ng/mL | 135.0 (84.8, 209.9) | 36.5 (19.3, 63.8) |
|  | Fibrinogen, mg/dL | 272.0 (240.0, 307.0) | 294.0 (263.0, 332.0) |
|  | Potassium, mmol/L | 3119.9 (2485.8, 3888.5) | 3208.5 (2563.7, 3988.4) |
|  | HGB, g/dL | 15.8 (15.2, 16.5) | 13.9 (13.3, 14.5) |
| Quantitative covariates* | Age, years | 39.8 (27.5, 51.3) | 39.5 (26.6, 51.4) |
|  | Physical activity, MET-min/week | 4068.0 (1953.0, 7146.0) | 2994.0 (1386.0, 5630.0) |
|  | BMI, kg/m^2^ | 1970.5 (1616.6, 2401.4) | 1827.0 (1489.2, 2222.3) |
|  | Total Energy Intake, kcal | 25.1 (23.1, 27.7) | 23.5 (21.4, 26.7) |
| Categorical covariates** | Smoking habit |  |  |
|  | *Never smoker* | 1478 (53.3) | 1770 (57.6) |
|  | *Current smoker* | 593 (21.4) | 590 (19.2) |
|  | *Past smoker* | 704 (25.4) | 715 (23.3) |
|  | Education |  |  |
|  | *Primary school or no title* | 112 (4.0) | 162 (5.3) |
|  | *Lower secondary school* | 348 (12.5) | 527 (17.1) |
|  | *Upper secondary school* | 228 (8.2) | 428 (13.9) |
|  | *Vocational school* | 611 (22.0) | 989 (32.2) |
|  | *University or higher* | 1476 (53.2) | 969 (31.5) |
|  | Following a special diet |  |  |
|  | *Yes* | 73 (2.6) | 145 (4.7) |
|  | *No* | 2702 (97.4) | 2930 (95.3) |
|  | Menstrual status |  |  |
|  | *Menstruation_Yes_* | --- | 832 (27.1) |
|  | *Menstruation_No_* | --- | 2243 (72.9) |

### **Table S5:** Table 1: Sex-stratified characteristics of the Healthy+ subsample

* median and interquartile range (IQR); ** N (%); ^ portions per week.

### **Table S6**: Results of the linear (LM) and generalized additive models (GAM) in the Healthy+ group. LM: Effects of DPs on eGFR in ml/min/1.73 m^2^ with 95% confidence intervals (CI) and p-values; GAM: Deviation from linearity (edf) and p-values.

|  |  |  | Linear model | | GAM | | |
| --- | --- | --- | --- | --- | --- | --- | --- |
|  |  |  | Coefficient (95% CI) | p.value |  | edf | p-value |
| Males  (N=2775) | | DASH | 0.67 (0.24, 1.09) | 0.00206 |  | 2.0 | 5.2E-03 |
|  |  | MDP_1_ | 0.83 (0.44, 1.21) | 0.00002 |  | 1.0 | 2.3E-05 |
|  |  | MDP_2_ | 0.14 (-0.32, 0.61) | 0.54240 |  | 2.3 | 1.4E-01 |
| Females  (N=3075) | | DASH | -0.12 (-0.49, 0.25) | 0.51815 |  | 1.7 | 0.5328 |
|  |  | FDP_1_ | 0.19 (-0.21, 0.6) | 0.34244 |  | 2.6 | 0.2215 |
|  |  | FDP_2_ | -0.39 (-0.73, -0.04) | 0.02784 |  | 1.0 | 0.0273 |
| Menstrual status | *Menstruation_Yes_* (N=2243) | DASH | -0.16 (-0.61, 0.28) | 0.47650 |  | 1.9 | 2.4E-01 |
|  |  | FDP_1_ | 0.09 (-0.39, 0.57) | 0.72307 |  | 1.3 | 9.2E-01 |
|  |  | FDP_2_ | -0.46 (-0.87, -0.05) | 0.02887 |  | 1.0 | 3.3E-02 |
|  | *Menstruation_No_*  (N=832) | DASH | 0.00 (-0.68, 0.67) | 0.99521 |  | 1.0 | 8.4E-01 |
|  |  | FDP_1_ | 0.68 (-0.07, 1.43) | 0.07748 |  | 1.0 | 3.5E-03 |
|  |  | FDP_2_ | -0.07 (-0.7, 0.57) | 0.83745 |  | 1.0 | 9.8E-01 |

### **Figure S1**: Analysis flowchart

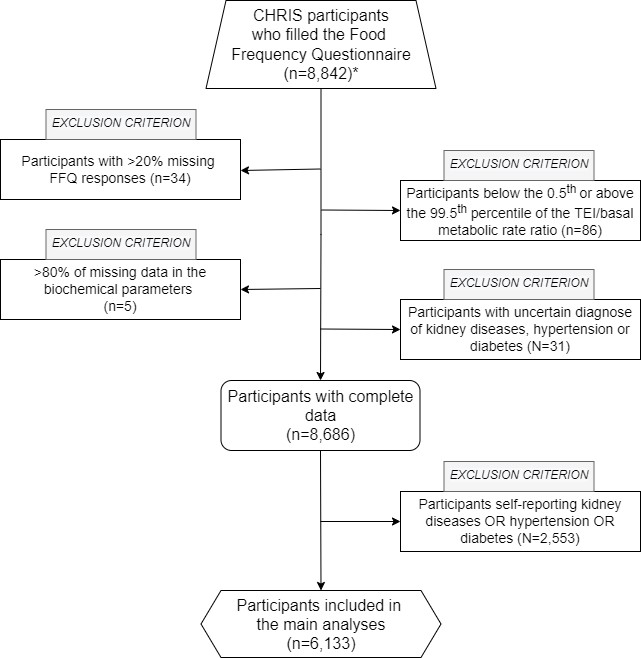

*Questionnaire was introduced starting from 2014-05-05

### **Figure S2**: Directed Acyclic Graph (DAG) used to identify confounders

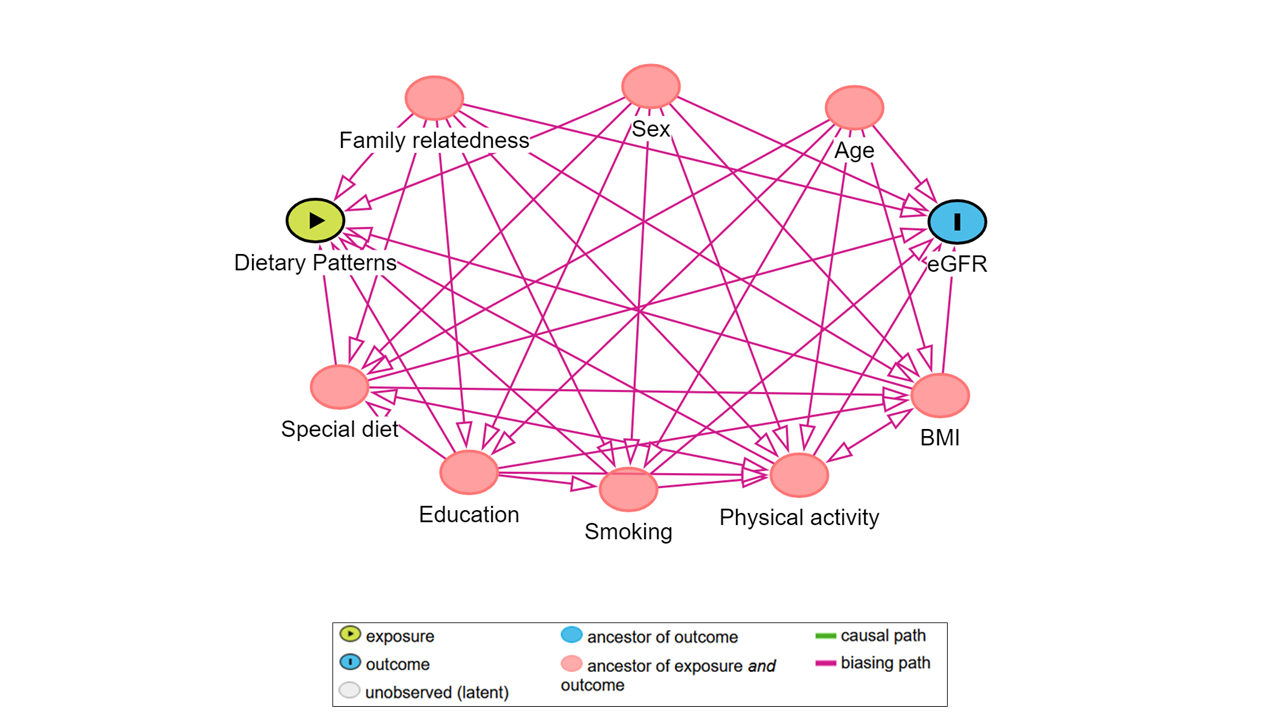
